# Prior knowledge informs graph neural networks to improve phenotype prediction from proteomics

**DOI:** 10.1101/2025.11.23.25340814

**Authors:** Prabuddha Ghosh Dastidar, Gus Fridell, Joshua M. Popp, Marios Arvanitis, Alexis Battle

## Abstract

High-throughput proteomics data provides dense individual-level molecular readouts, enabling the development of machine learning models for predicting diverse phenotypes relevant to patient health. Proteins interact in the cell in complex, nonlinear relationships that may not be reflected by linear models or simple machine learning approaches, highlighting the potential for more expressive deep neural networks to improve performance. Despite this possibility, in practice, developing neural network approaches in biological domains has been a significant challenge. We developed a deep learning framework for predicting disease-related traits from protein expression data using an innovative model architecture designed to exploit structured biological knowledge. The core of the model is a graph neural network (GNN) operating on bipartite graphs where one set of nodes represents protein expression levels and the other represents hundreds of protein sets derived from gene ontology libraries. Edges encode set membership, providing a compact and biologically meaningful structure. We trained our model using the UK Biobank plasma proteomics and individual phenotype data. Of the architectures we examined, the best-performing architecture had three parallel heads: two GNNs each using graphs constructed with independent protein set libraries and one global head consisting of tabular protein expression data. Their outputs are concatenated and passed through a dense feed-forward network to predict phenotype. When applied to predicting glycated hemoglobin (HbA1c) levels and a range of other phenotypes, our model showed strong predictive performance, outperforming other deep learning architectures and simpler linear models. Control models with permuted protein labels displayed worse performance demonstrating that the model benefits from the inductive bias from incorporating prior knowledge, especially in settings with limited training data. We present an innovative model architecture incorporating biological domain knowledge to predict complex traits from large scale proteomic data.

## Introduction

The growing availability of deep molecular profiling creates new opportunities for precision medicine and personalized disease risk prediction from data such as proteomic measurements.^1^ Proteins form the machinery of the cell, and ultimately mediate much of the relationship between genetics and disease phenotypes, making proteomics a particularly attractive data type for translational applications.^2^ Large biobank data offer the potential to develop and train predictive models that capture the impact of protein levels on disease-related phenotypes. The biological mechanisms reflected by protein levels may include complex, nonlinear interactions that are often missed by linear models or simple machine learning approaches, indicating an opportunity for improvement through application of deep neural networks.^3^ However, standard deep learning architectures tend to require very large sample sizes and are prone to poor performance in data-limited settings^4^. Indeed, the sample sizes available with paired molecular and phenotype information, even in large biobanks of tens of thousands of individuals, may not be sufficient alone for training large models. These limitations highlight the need for architectures that incorporate domain knowledge to constrain the hypothesis space in ways that align with biological structure.

A growing body of work suggests that introducing biologically motivated inductive biases into neural networks can improve model performance.^5–7^ Biologically-informed, structured models constrain the hypothesis space, which may reduce overfitting and therefore enable improved generalization and higher predictive accuracy.^8^ Recent work has demonstrated that thoughtfully incorporating domain knowledge into deep learning models based on biological pathways can offer improvements such as increasing power in genome wide association studies^5^ and accurately simulating cell growth^6^. Here, we focus on methods for incorporating domain knowledge in model architectures to improve performance of models predicting phenotype from proteomics. Identifying useful forms of inductive bias that are effective for large-scale proteomic prediction and determining how these should be incorporated into neural network architectures remains an open question.

We introduce a deep learning framework designed to address this challenge. We embed structured prior biological knowledge, represented in the form of a graph capturing protein co-functionality, directly into the model architecture. The core of the model is a graph neural network (GNN) operating on bipartite graphs where one partition of nodes represents proteins and the other represents protein sets derived from Gene Ontology (GO)^9,10^ libraries reflecting current knowledge of the functional relationships between genes and their membership in multiple cellular processes. Trained on proteomics from the UK Biobank Pharma Proteomics Project (UKB-PPP)^11,12^ and UK Biobank (UKB) phenotyping data, this architecture allows the model to learn predictive signals while incorporating known molecular function and cellular context, demonstrating better generalization in settings with limited training data and improving interpretability. Benchmarking against baseline models without biologically-informed architectures demonstrates that our model benefits from the inductive bias imposed through graph structure. Our model is a modular GNN framework that leverages information from curated gene sets (used in the manuscript interchangeably with “protein sets” as constructed by mapping each gene to its corresponding protein product) to more effectively learn from high-dimensional proteomic data for phenotype prediction.

## Methods

### Graph construction and informed GNN architecture

#### Construction of protein graphs for use in GNNs

We constructed bipartite graphs to integrate prior biological knowledge into the model architecture. The bipartite graph contains two node types. One partition of nodes represents individual proteins, which will be embedded with standardized protein expression values derived from the UK Biobank proteomic dataset for each instance (individual). The other partition represents protein sets, in our case curated from GO libraries, although other resources could be utilized. Edges encode membership relationships between proteins and their associated protein sets, effectively linking measured protein features to higher-level biological processes. Each protein set node is initialized with a constant scalar rather than learned parameters, ensuring that any downstream signal propagation reflects relational structure rather than initial feature weighting. This design imposes functional relationships into the network structure itself, serving as an inductive bias that constrains learning to biologically meaningful dependencies. This approach also helps regularize learning in high-dimensional proteomic spaces by reducing the model’s hypothesis space compared to a message passing scheme with no prior knowledge.

#### Single-head GNN architecture

Our basic framework is built on a GNN architecture. We first introduce the approach constructed using a single bipartite graph, where protein nodes and protein set nodes are connected through the GO-derived membership structure. Here, we used the GO Molecular Function library. The model consists of two stacked graph convolutional network (GCN) layers (with dimensions 1 to 2 and 2 to 1), each using the bipartite graph to aggregate messages between protein and protein set nodes. Self-loops in the graph are added to ensure consideration of unconnected protein nodes. Both layers used hyperbolic tangent activation functions to maintain a bounded latent representation and prevent node zeroing. Having two GCN layers allows communication between nodes separated by up to two edges. Consequently, proteins with shared set membership communicate with each other. These layers enable bidirectional information flow between proteins and their associated sets, enriching protein representations with higher-level functional context. After GCN layers, the updated protein node embeddings are concatenated to form a single vector of protein embeddings for each individual.

This graph-derived embedding is then concatenated with the corresponding individual scalar covariates (see “Training Scheme and Model Specifics” section below for specific covariates included) and passed through a fully connected network consisting of four layers with dimensions as follows: double the dimension of the graph-derived embedding vector (one entry per protein) plus number of additional inputs (e.g. covariates), 256, 32, and 1 (final prediction). The first three dense layers use LeakyReLU activations with a negative slope of 0.01, while the final layer is linear and outputs the predicted continuous phenotype value. Training uses mean squared error loss, the Adam optimizer, and adaptive learning rate reduction with early stopping for regularization. Model hyperparameters (e.g. learning rate, dropout rate) are selected through validation performance. Refer to the “Training Scheme and Model Specifics” section for the training scheme and packages used. The GCN layers and the dense feed-forward prediction head were trained in series, end-to-end.

In summary, graph-derived protein embeddings are generated through two layers of graph convolution, which propagate and integrate information across protein set connections. The resulting protein-level embeddings are flattened to form an individual-specific vector representation. Individual covariates are then concatenated with these embeddings. This combined representation is passed through a fully connected network to predict the outcome of interest. This architecture allows the model to propagate information between proteins that have biological similarity, leveraging ontological knowledge.

#### Parallel GNN architecture

Building on the single-head design, the parallel dual-graph model (Figure 1) was developed to extract refined protein embeddings that are specific to different GO ontologies while mitigating the risk of over-smoothing by combining this information into a single, densely connected graph. The architecture is the same as the single-head model except the input graph is different. Instead of one bipartite graph, the input graph contains two independent bipartite subgraphs. The model operates on the two independent bipartite subgraphs in parallel with no edges between them. The protein node partition is duplicated to be the same in each subgraph, but it connects to a distinct set of gene set nodes derived from a different GO library. One subgraph encodes Molecular Function relationships, while the other encodes Cellular Component associations. This produces two ontology-specific protein embeddings that capture distinct contextual information. This separation enables the model to learn complementary aspects of protein activity, functional role, and cellular localization, without forcing these representations to converge through shared message passing. The embeddings from both subgraphs are then concatenated to form a unified vector representation for each individual, combining signals from both GO libraries while preserving the individuality of each source.

**Figure 1.**
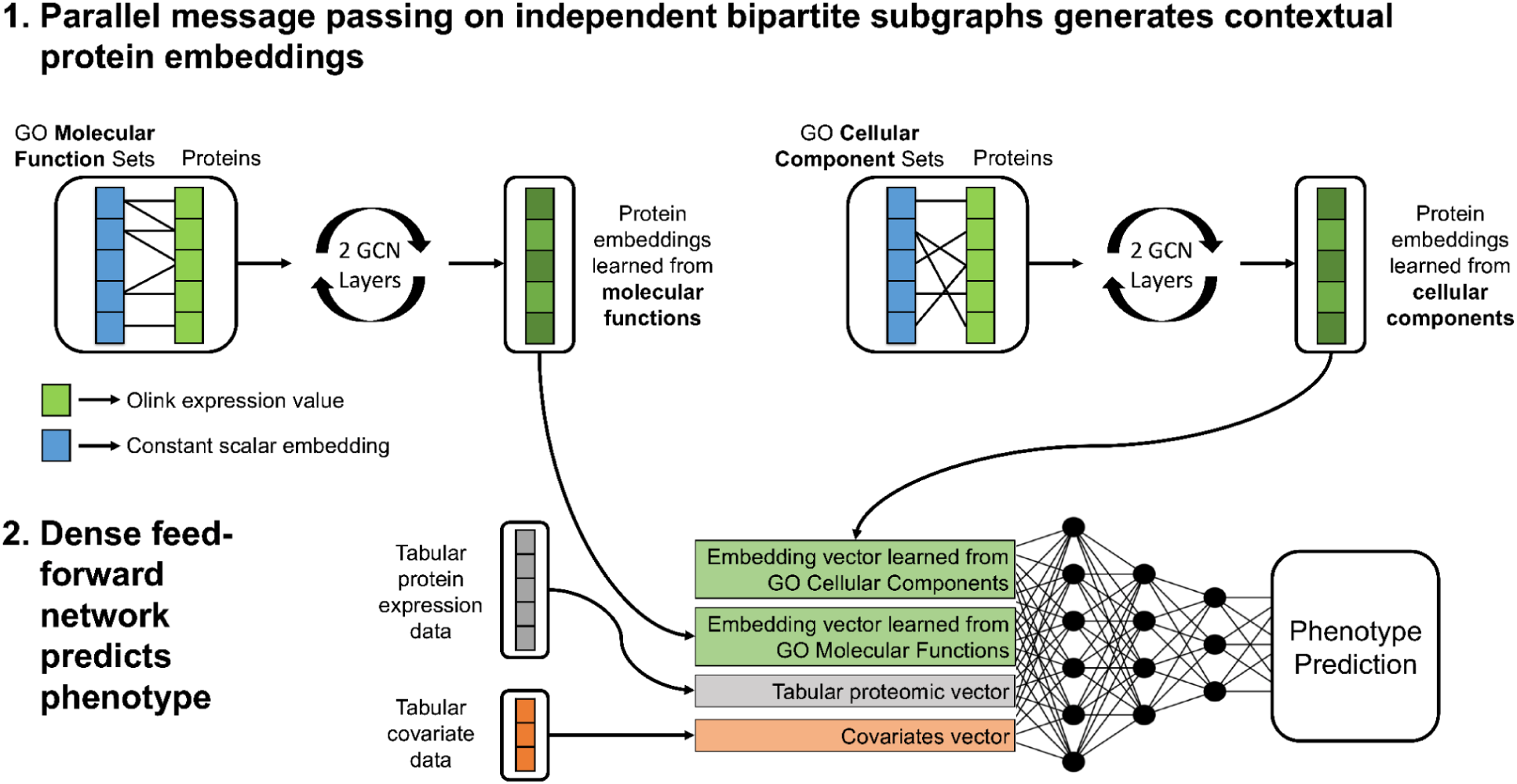
Overview of the parallel graph neural network architecture for proteomic phenotype prediction. The model operates on two independent bipartite subgraphs derived from GO libraries. In each subgraph, protein nodes (green) are connected to GO set nodes (blue) representing either Molecular Function or Cellular Component relationships. Protein nodes are initialized with expression values, while set nodes are initialized with constants, allowing two graph convolutional (GCN) layers to propagate contextual information between proteins that share set membership. The resulting ontology-specific protein embeddings are concatenated with raw proteomic features and subject covariates before entering a fully connected feed-forward network with three hidden layers to predict the phenotype. The GCN layers and the dense feed-forward prediction head were trained in series, end-to-end.

Downstream layers and training procedures are identical to those described for the single-head GNN. The concatenated embedding is augmented with individual-level covariates and the unchanged tabular proteomic data, then passed through a fully connected feed-forward network. The introduction of unchanged tabular proteomic data as an additional input right before the feed-forward network serves as a global head, exposing the model to the individual protein levels in addition to the GNN embeddings.

By maintaining separate message-passing channels for each ontology, the parallel dual-head architecture aims to reduce oversmoothing and allows proteins to adopt differentiated embeddings across biological contexts.

#### GNN feature interpretation

For the single-head GNN, we quantified the importance of each protein set using a gradient-based saliency approach. For each test graph, we enabled gradients on the input node features and backpropagated the loss to compute how much each feature influenced the model output. We then extracted the gradients corresponding to the protein and protein set nodes and took their absolute values as saliency scores. These scores were averaged across all test graphs to obtain a single importance value per protein set, which was used for ranking and further analysis.

### Baseline and random graph models

#### Baseline models

##### Permuted GNN

To isolate the effect of prior knowledge from GNN structure on model performance we also trained permuted GNNs. In these models, the bipartite graph connectivity was randomly permuted while maintaining node degree distributions and embedding dimensions. This was done by simply permuting protein names prior to the graph construction process. This destroyed the correspondence between proteins and their true functional sets while preserving graph topology and model capacity. Thus, differences in predictive performance between the true-graph GNN and the permuted GNN isolate the benefit of prior knowledge from general model capacity. For a given model, its permuted baseline refers to the same model architecture with protein names being permuted before graph construction.

##### Large single-head GNN

To provide a baseline for the parallel dual-graph GNN, a large, connected-graph single-head GNN was implemented. This model encodes information from both GO Molecular Function and GO Cellular Component libraries within a single bipartite graph, rather than in two independent subgraphs. The protein partition remains the same, but the gene set partition merges all sets from both ontologies, creating a single, more densely connected graph. The rest of the architecture and training procedures are identical to those described for the parallel GNN. This configuration tests whether integrating information from multiple ontologies into a shared message-passing space leads to oversmoothing and loss of representational specificity, compared to the refined, ontology-specific embeddings learned in the parallel GNN.

##### Informed feed-forward network

This baseline model was a feed-forward neural network with protein expression levels as input without any GNN component. The first hidden layer contains one node per gene set from Gene Ontology Molecular Functions. These set nodes are only connected to proteins in the input layer that belong to each set. The following four hidden layers were dense with 256, 128, 64, and 32 nodes, respectively. Batch normalization and dropout were applied to all layers to improve stability and prevent overfitting. The first layer used a hyperbolic tangent activation, while subsequent layers used LeakyReLU activations. The model was trained using the Adam optimizer with a learning rate scheduler and early stopping based on validation loss.

##### Ridge regression

As a linear benchmark, ridge regression was trained on the proteomic data to predict phenotype, selecting the regularization strength through validation set performance.

#### Random graphs GNN

To isolate the effect of model size on performance, we created the random graph GNN. The random graph GNN follows the same structure as the parallel architecture, including independent bipartite subgraphs processed in parallel and identical downstream feedforward and training procedures. The key difference is that the edges are not biologically derived. Instead of ontology-based gene sets, random set libraries are used to define the bipartite graphs. A random set library consists of 200 randomly generated sets, where each set includes 2-20 (size for each set randomly sampled from uniform distribution) proteins chosen uniformly at random.

Five separate models were trained, each containing 1 to 5 independent bipartite subgraphs (random graph heads), corresponding to different random set libraries. As in the parallel ontology-based GNN, this concatenated embedding is then combined with a global head prior to input into the feed-forward network. The global head consists of the unchanged tabular proteomic data and individual-level covariates.

#### Random single-head GNN

To evaluate whether oversmoothing influenced model performance, independent of ontological structure, a random single-head GNN was trained using the same architecture and training configuration as the single-head ontology-based model. The only difference was the graph structure: instead of GO-derived gene sets, the model operated on a single bipartite graph constructed from randomly generated protein sets.

The number of sets in the graph was varied from 100 to 1000 in increments of 100, allowing systematic assessment of how graph density affects learning and potential oversmoothing. The sizes of the random sets were sampled from a geometric distribution parameterized to mimic the empirical distribution of set sizes observed in the GO Cellular Component library. Protein memberships of each set were assigned uniformly at random.

### Experimental setup and training procedures

#### Model comparison

To systematically assess the contribution of graph structure and ontology-specific design choices, several controlled comparisons were performed:

##### Single-head GNN vs. informed FFNN

This comparison isolated the impact of the GNN architecture.

##### Single-head and parallel GNN vs. permuted versions

For each ontology-based GNN, a permuted control was constructed by shuffling protein identities prior to graph construction. This maintained graph degree distributions and model architecture while destroying true biological relationships in the true graph, isolating the predictive value of inductive bias through meaningful graph structure. We trained on 25%, 50%, and 100% of the training data to evaluate the relationship between inductive bias in the model and data availability.

##### Parallel GNN vs. large single-head GNN

This comparison evaluated the effect of keeping ontology-specific embeddings separate. In the Large Single-Head model, Molecular Function and Cellular Component sets were combined into one bipartite graph, testing whether the parallel design helps mitigate oversmoothing by maintaining independent message-passing spaces.

##### Random graph GNNs

These models followed the same structure as the parallel GNN but used between one and five independent bipartite subgraphs built from randomly generated protein sets. This controlled for model size independent of biological inductive bias. We trained on 25%, 50%, and 100% of the training data to evaluate the relationship between model size and data availability.

##### Random single-head GNN

To further test oversmoothing, single-head models were trained on random bipartite graphs where the number of sets varied from 100 to 1000. This assessed the effect of increasing graph density and connectivity independent of biological inductive bias.

#### UK Biobank proteomic data and preprocessing

All analyses were conducted using proteomic and phenotype data from the UK Biobank Pharma Proteomics Project (UKB-PPP). To minimize batch and cohort-related confounding, participants selected by the consortium members (based on specific disease or population characteristics) and participants selected for a COVID-19 study were removed from the dataset. Participants with more than 500 missing protein measurements and proteins missing in more than 2000 participants were removed. Remaining missing values were mean-imputed using the average expression of each protein across all remaining participants. Individuals missing measurements for the phenotype being predicted were removed from the dataset. The filtered dataset (for results shown in the main body) contained nearly 37,000 participants prior to data splitting. The phenotype analyzed in the main body of the study was glycated hemoglobin (HbA1c; UK Biobank field ID 30750). Analysis of additional phenotypes are in the supplemental figures. The additional phenotypes are blood C-reactive protein (CRP) measured from serum biochemistry (UKBB Field 30710), pack years smoked (UKBB Field 20161), and standing height (UKBB Field 50).

#### Training scheme and model specifics

Data were divided into 75% training and 25% testing subsets, with 15% of the training data reserved as a validation set for hyperparameter (learning rate and dropout rate) selection. The best performing model (see “Parallel GNN architecture” section above), when trained on 100% of the training data, had a learning rate of 0.00001 and a dropout rate of 0.5. Model optimization used the Adam optimizer with mean squared error loss, early stopping, and an adaptive learning rate scheduler that halved the learning rate after five epochs without validation improvement. Graph structures were constructed from the GO Molecular Function and Cellular Component libraries, including only sets containing between 2 and 100 proteins present in the final dataset. Covariates incorporated as scalar inputs included sex, age, volume of Li-heparin plasma in the UKB sample (technical covariate), genomic principal components 1–16 (ancestry), and body mass index (BMI). Packages used for graph neural networks, feed-forward neural networks, ridge regression, and gene set construction were PyTorch Geometric^13^, PyTorch^14^, scikit-learn^15^, and Enrichr^16–18^, respectively.

## Results

### Incorporating biological knowledge into GNN model architecture improves predictive performance

We tested the single-head GNN to isolate the effect of biologically-informed architectures on phenotype prediction. Here, we focus on the prediction of HbA1c due to large sample size and association with numerous proteins. Additional phenotypes including blood C-reactive protein (CRP) measured from serum biochemistry (UKBB Field 30710), pack years smoked (UKBB Field 20161), and standing height (UKBB Field 50) were also evaluated as supplementary support for our conclusions. When imposing domain knowledge through graph structure derived from just GO Molecular Function library sets, model performance consistently improved relative to permuted baselines (green bars, Figure 2A, 2B and Supplementary Figure 1, 2). As many proteomic datasets are comprised of fewer samples, we examined the impact of training data size on model performance. Performance, when predicting HbA1c, decreased significantly as the training set size was reduced to 50% (11,715 samples) or 25% (5858 samples), though the model still performed above the permuted baseline in each case. This demonstrates that the inductive bias provided by biologically meaningful graph structure improves the models’ ability to utilize signal from high-dimensional proteomic data and this result holds in settings with limited training data. This suggests that imposing curated biological structure into the model architecture acts as a regularizer, constraining learning to functionally plausible relationships among proteins. In low-data settings especially, such structure effectively reduces the search space of possible mappings between protein expression and phenotype, helping the model generalize better.

**Figure 2.**
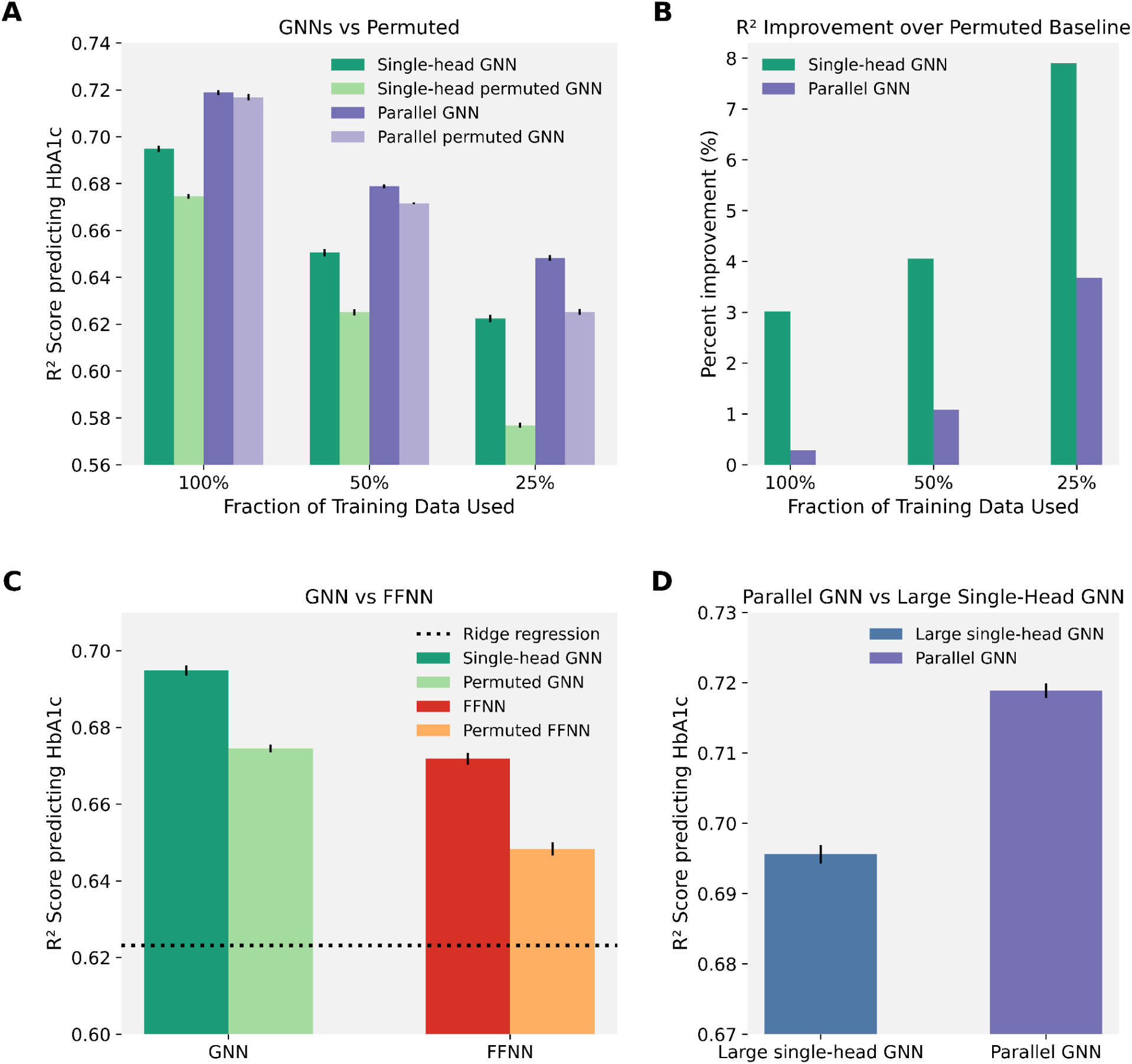
Comparative evaluation of graph neural network architectures and baseline models for HbA1c prediction. **(A)** *Informed GNNs vs. permuted graphs.* The single-head and parallel GNNs, which operated on the protein graph derived from GO gene sets, were trained on the full training data (100%) and smaller subsets of it (50%, 25%). Each model was compared to a corresponding permuted-graph baseline (see “Baseline models” section in Methods) in which protein names were permuted to remove biological meaning from the graph. Bars represent mean test R^2^ across 10 independent training runs (± S.E.M.). **(B)** *Improvement over permuted baselines.* Mean percent improvement in R^2^ (represented by bars) was computed for each model in (A) relative to its permuted-graph baseline (see “Baseline models” section in Methods), based on 10 independent training runs (with varying seeds). **(C)** *GNN vs. feed-forward neural network baseline.* The single-head, ontology-informed GNN was compared to an ontology-informed, feed-forward neural network baseline (see “Baseline models” section in Methods) trained on the same samples. Both models were also evaluated against their respective permuted baselines where protein names were permuted. Bars indicate mean R^2^ across 10 independent runs (± S.E.M.). The black dotted line indicates the test R^2^ when using ridge regression to predict HbA1c. All shown metrics are from training on 100% of the training set. Test R^2^ for ridge regression when training on 50% and 25% of the training data (not shown) were 0.5831 and 0.5320, respectively. **(D)** *Parallel vs. large single-head GNN.* The parallel GNN was compared to a large single-head GNN baseline (see “Baseline models” section in Methods). Bars show mean test R^2^ over 10 independent training runs (± S.E.M.).

### Parallel GNNs incorporating multiple views of protein relationships outperforms single-head GNNs

The benefit of inductive bias through building the model architecture with prior knowledge is clear when we build the graph using GO Molecular Function library sets in the single-head GNN. However, we wanted our graph structure to incorporate information from multiple informative GO gene sets, including Molecular Function and Cell Component. The most direct approach is to include both libraries in the graph structure of the single-head GNN, which is what we did in the large single-head GNN baseline. In contrast, the parallel GNN, which incorporated information from two GO set libraries through parallel message passing operating on two separate graphs, preserved separation between molecular and cellular contexts, allowing each graph to encode complementary aspects of protein biology before integration of both embeddings for the prediction task. The parallel GNN outperformed the large single-head GNN baseline which contained two ontology libraries in one graph (Figure 2D). We hypothesize that the large single-head GNN led to oversmoothing or “greying out” signal, which hindered performance. The parallel GNN achieving the best performance indicates that biologically motivated modularity, rather than naive graph expansion, is key to effective inductive design in proteomics GNNs.

Additionally, among all architectures tested, the parallel GNN model yielded the best predictive performance on phenotype. First, it outperformed the single-head GNN architecture (with information from GO Molecular Function only. This single-head GNN architecture outperformed a traditional feed-forward neural network and the linear baseline (ridge regression) when predicting HbA1c levels (Figure 2A, 2C). The performance boost from the parallel GNN compared to the single-head GNN (with information from a single set library) is likely due to the incorporation of more domain knowledge (through GO Cellular Component sets) and the increased model size. In the traditional feed-forward network, manually wiring the early network layers to reflect protein pathway membership guided the model toward biologically meaningful prediction since this model outperformed its permuted counterpart (Figure 2C). However, this model left learning higher-order interactions between GO sets to fully unconstrained portions of the network, which is why we hypothesize it was outperformed by our GNN framework, which explicitly encodes interpretable protein set membership structure into the architecture and allows the model to refine individual protein expression embeddings based on this ontological context.

### The benefit of inductive bias in GNN architectures is more pronounced in settings with limited training data and model size

In both the parallel and the single-head GNN, prior knowledge in the form of gene sets boosted performance as these models outperformed their permuted counterparts. However, the relative benefit of this inductive bias diminished as the amount of training data increased (Figure 2B, Supplementary Figure 2). Demonstrating the benefit of its inductive bias, the parallel GNN outperformed its associated permuted baseline when models were trained on 50% and 25% of the training data. However, with full training data the parallel GNN performed nearly identically to its permuted counterpart (Figure 2A, 2B). Notably, however, the single-head GNN outperformed its permuted baseline for all training data fractions, demonstrating the importance of inductive bias in smaller, lower parameter models. With larger training datasets, the permuted models without biologically meaningful graphs approached similar performance to the ontologically-informed models, suggesting that data volume can partially compensate for the lack of structural prior knowledge. As the training data expands, it is likely that the model becomes increasingly capable of inferring dependencies from the data itself, which explains the diminishing marginal benefit of the prior knowledge. This suggests that a model large enough and with enough training data may possibly compensate for a lack of inductive bias in the architecture. Though, inductive bias in low sample settings and smaller models considerably improves predictive performance.

### Feature importance on single-head GNN reveals relevant biology

Feature importance (Figure 3) analyses on the single-head GNN showed that the model prioritizes biologically meaningful protein sets when predicting HbA1c, a clinical marker of diabetes. This is particularly notable since GO set nodes in the graph, unlike the protein nodes, were not directly used as input nodes for the dense network; they served as moderators of protein interaction and were all initialized with the same constant scalar. Yet, feature importance tests on these GO set nodes still detected relevant biology. Among the most influential sets were those related to dipeptidyl peptidase activity, which includes DPP4, GLP1, and GIP and corresponds to established drug targets for diabetes, as well as fatty acid binding.^19,20^ For the protein nodes, the gradient saliency analysis also highlighted proteins, like SHBG, with potential associations with HbA1c (and its associated phenotypes).^21,22^ These results show that the model potentially captures disease-relevant structure.

**Figure 3.**
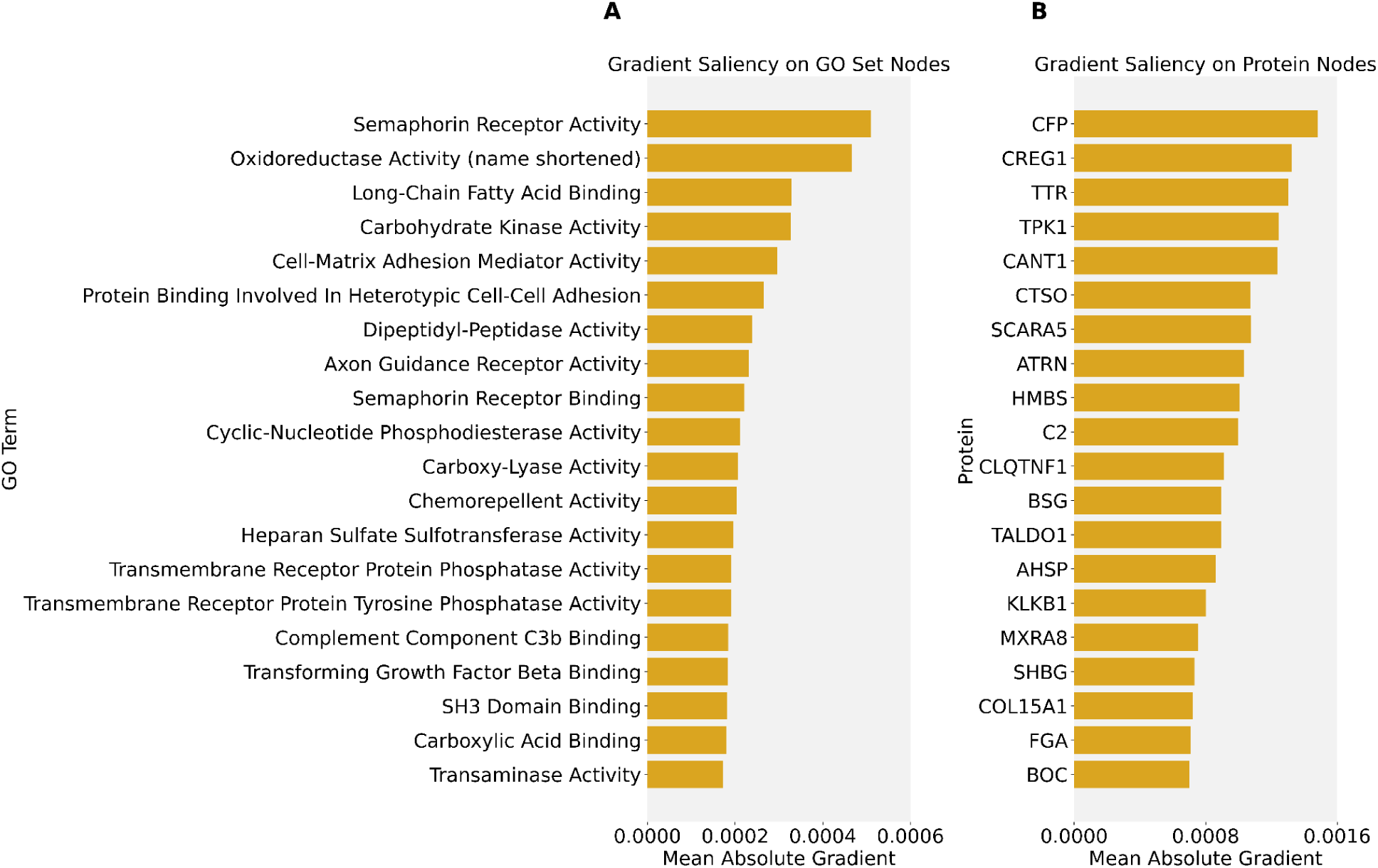
Gradient-saliency based feature importance scores for GO set nodes and protein nodes. Mean absolute gradient scores are shown for the top 20 **(A)** GO term nodes and **(B)** protein nodes, computed across all test samples. Bars represent the magnitude of the mean gradient with respect to each node. GO terms used are from GO Molecular Function.

### Increasing model size boosts performance under sufficient training data

There is improvement seen from the parallel GNN (incorporating information from two GO libraries) over the single-head GNN (which only incorporated information from GO Molecular Function), and this could be attributed to two factors: the incorporation of extra GO knowledge in the model architecture and the increased model size that comes from adding additional heads. To disentangle the effects of larger model size from inductive bias, we compared models of varying depth and complexity with a random graph GNN. We repeated all tests on three different training data fractions: 100%, 50%, and 25%. We first found that simply increasing the number of random graph heads led to better predictive performance (Figure 4A and Supplementary Figure 3). However, we identify a trade-off that exists between model size and training data availability. When predicting HbA1c levels, the number of random graph heads required to reach peak performance decreases as the model trains on smaller datasets. This indicates diminishing returns when training data is limited: we hypothesize that the larger random graph GNN models can, in principle, learn a larger range of relationships and increasingly complex relationships, but only when supported by sufficient data. The reduction in required heads in smaller data regimes underscores that careful architectural design (achieved through structured prior knowledge) can sometimes work better than brute-force model enlargement when data is limited. Particularly, inductive bias and model capacity (determined by its size) play complementary roles: the former constrains the learning space, while the latter expands it, and the optimal balance changes depending on data availability.

**Figure 4.**
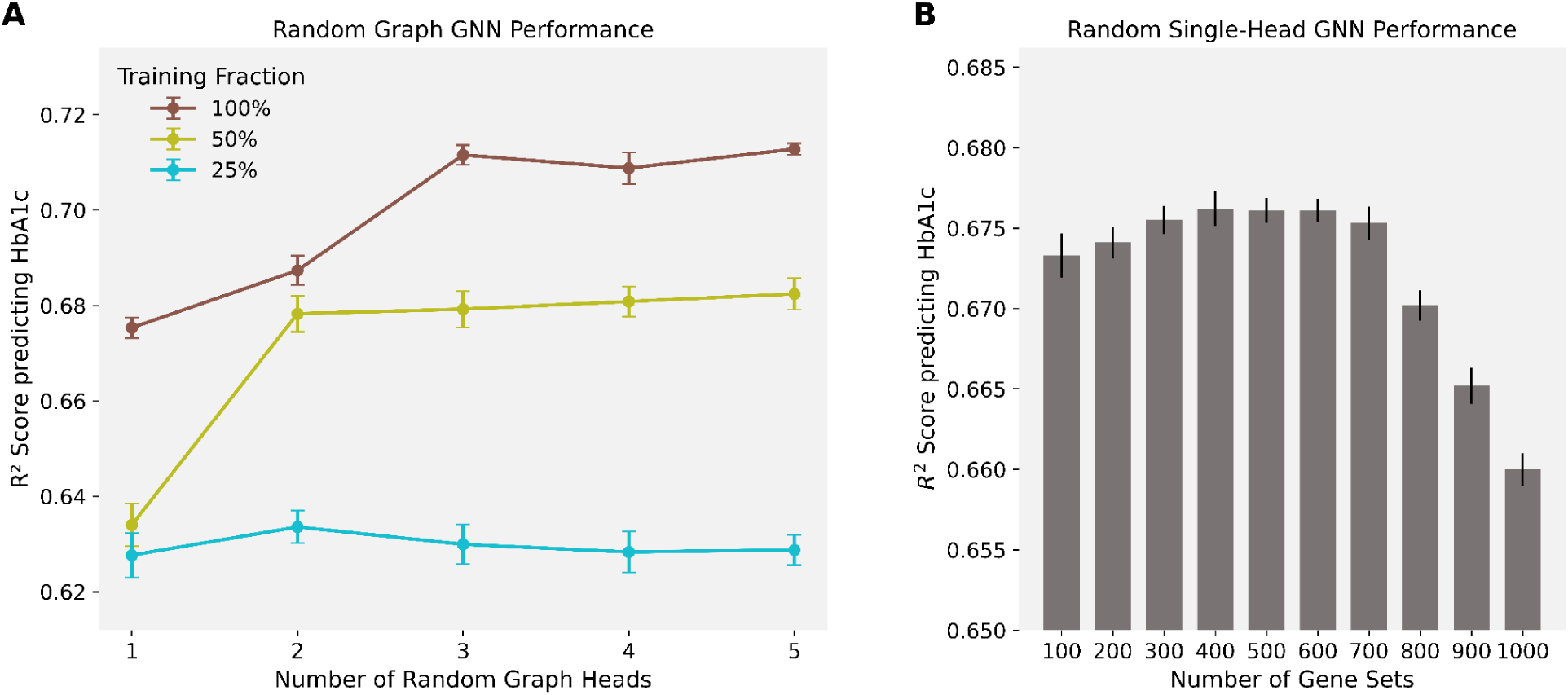
Performance of GNNs trained on random graph structures. **(A)** *Random graph GNN performance.* GNNs were trained using randomly generated bipartite graphs containing between one and five random graph heads, with 100%, 50%, and 25% of the training data used for each configuration. Points represent mean test R^2^ across 10 independent training runs (± S.E.M.). **(B)** *Random single-head GNN performance.* Single-head GNNs were trained on randomly generated bipartite graphs containing between 100 and 1000 random protein set nodes. Bars show mean test R^2^ over 10 independent training runs (± S.E.M.).

Attempting to increase model capacity through graph expansion must be balanced against the risk of oversmoothing. We observed that with a random single-head GNN, enlarging the random gene set library with more random gene sets (thereby expanding model size by increasing graph density instead of adding parallel random graph heads) can degrade performance (Figure 4B). When too many random connections are introduced, message passing might cause node embeddings to become homogenized, reducing their discriminative power. We hypothesize that beyond a certain point, adding complexity through denser graphs does not enhance the model’s capacity to learn more complex associations but instead blurs meaningful feature distinctions, reinforcing the importance of maintaining structured sparsity in graph-based models.

## Discussion

Incorporating structured prior knowledge improves phenotype prediction from proteomic data, with both the single-head and parallel GNN architectures outperforming their respective permuted baselines across multiple phenotypes. The magnitude of this improvement decreases as more training data is available, indicating that the benefit of domain-informed inductive bias is strongest under data-limited conditions. Notably, the parallel GNN, which provides embeddings influenced by two separate protein graphs in addition to individual raw protein levels, achieves the strongest overall performance across all training fractions. Even though its absolute accuracy is lower, the single-head GNN shows the largest relative gain over its permuted baseline. The parallel GNN model’s smaller relative advantage reflects that its added capacity may compensate for a lack of inductive bias in settings with high sample size. The random graph GNN reflects how, independent from the effect of informing the model architecture with prior domain knowledge, increased model capacity (which comes from expanding the model size) becomes more advantageous only when sufficient data is available to support it. These trends collectively highlight that inductive biases enhance GNN performance especially in low-data regimes, and that increasing model size and complexity may partially compensate for the lack of inductive bias as data abundance increases.

By building models that encode biological domain knowledge directly into their structure, we can extract more signal from high-dimensional proteomic data. More broadly, this demonstrates that model architecture itself can serve as a form of knowledge representation—encoding biological structure directly into the learning process rather than relying solely on the data. This approach aligns with the emerging paradigm of knowledge-guided machine learning, which seeks to combine the interpretability and generalizability of mechanistic models with the flexibility of deep learning. In the context of proteomics, such architectures may ultimately facilitate discovery by identifying which structured relationships are most predictive of phenotypic outcomes, offering a path toward biologically interpretable deep learning schemes.

There are some key limitations to this work. Because the proteomic measurements were obtained from plasma, our ability to draw context-specific biological conclusions is limited; plasma contains proteins released into circulation from tissues throughout the body.^23^ At the same time, this broad origin means the dataset captures signatures of many physiological processes. Additionally, for certain phenotypes, including height, the deep learning architectures evaluated do not exceed the performance of linear baselines (Supplementary Figure 1), suggesting that the advantages of ontology-informed models may depend on the degree of nonlinearity in the underlying trait architecture. Moreover, the graph representation used in the informed GNNs relies on protein modules defined by GO annotations, which may omit biologically meaningful groupings that are not well captured by GO libraries.

Future work could take advantage of other resources for prior knowledge and further investigation of architectural choices. For example, we extend the current GNN framework to better capture the hierarchical organization of biological knowledge and improve interpretability. A hierarchical graph connecting proteins to multiple ontology levels could enable the model to propagate information both within and across levels. In another direction, incorporating a multi-head graph attention mechanism could further enhance interpretability by learning adaptive weights for protein set connections, allowing identification of proteins, GO terms, and cross-level relationships that are most influential for specific predictions. Finally, other sources of information such as other gene sets and annotations of individual proteins may further improve performance. For example, protein nodes could be initialized with their expression values along with additional biological information, such as features derived from protein sequences^24^ or genetic prioritization scores^25^.

Overall, biologically-informed GNN architectures provide a powerful and interpretable framework for modeling high-dimensional proteomic data. Incorporating structured biological prior information into model design enables improved phenotype prediction, particularly in data-limited settings, while also highlighting relevant molecular pathways and functional relationships. These findings demonstrate that deep learning models can be both expressive and biologically meaningful when guided by curated knowledge, offering a promising avenue for translational proteomics and precision medicine applications.

## Data Availability

Data are available upon application to the UK Biobank: https://www.ukbiobank.ac.uk/use-our-data/apply-for-access/

## Acknowledgements

We would like to thank Rebecca Keener, Eduarda Vaz, and other members of the Battle lab for helpful discussions. This research has been conducted using the UK Biobank Resource under Application Number 98985. A.B. was supported by NIGMS R35GM139580.

## Author contributions

P.G. participated in project conceptualization, formal analysis, investigation, methodology, data curation/visualization, software, and writing. G.F. participated in project conceptualization, methodology, administration, supervision, writing, and code review. J.P. participated in project conceptualization, methodology, administration, and supervision. M.A. participated in validation. A.B. participated in project conceptualization, administration, supervision, funding acquisition, and writing.

## Declaration of interests

A.B. is a shareholder in Alphabet Inc, and a founder and equity holder of CellCipher, Inc. J.P. is an equity holder of CellCipher, Inc.

---

**Supplementary Figure 1.**
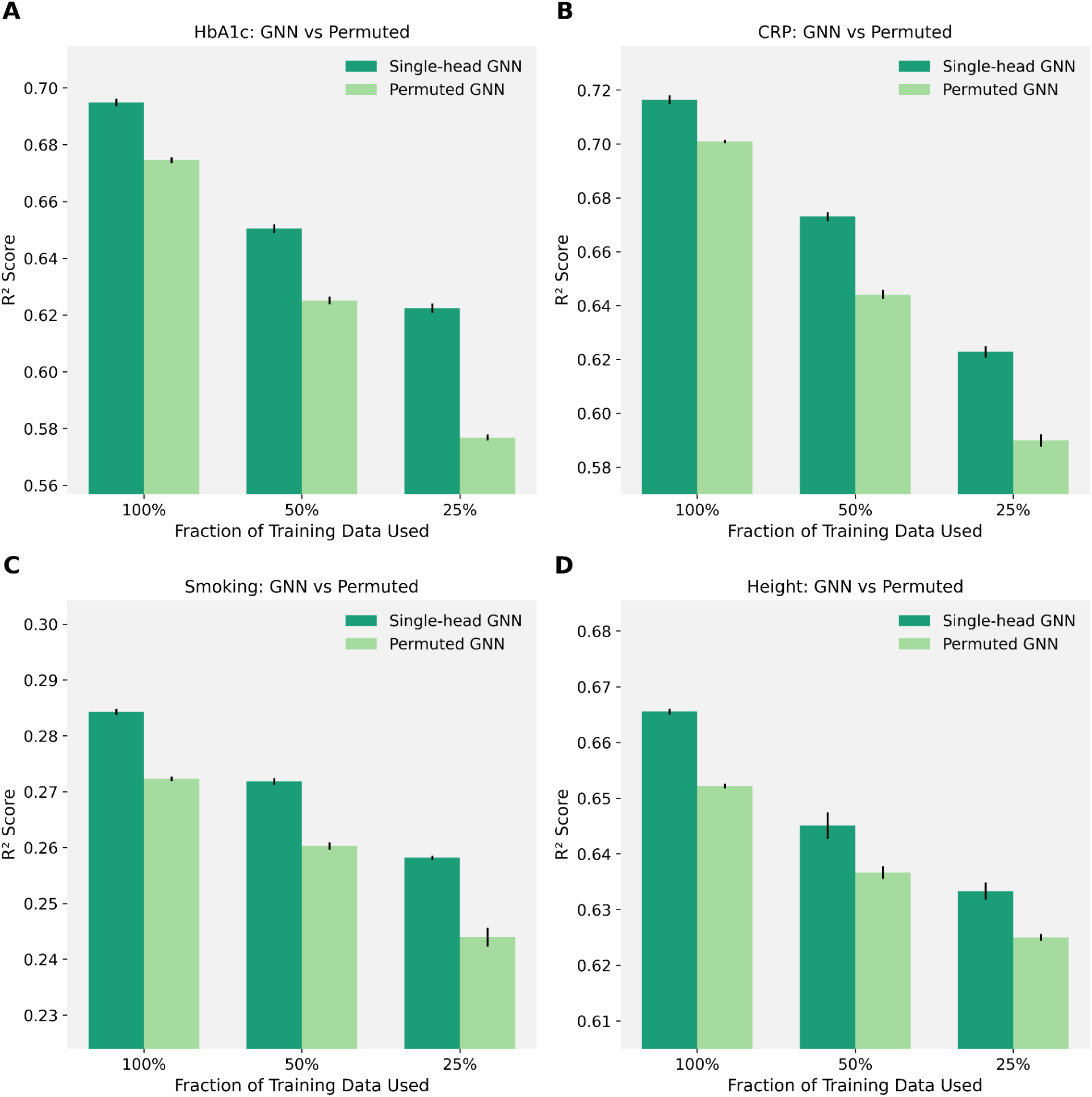
Comparative evaluation of single-head graph neural network architectures and baseline models for multiple phenotype prediction. The single-head GNNs, which operated on the protein graph, were trained on the full training data (100%) and smaller subsets of it (50%, 25%). Each model was compared to a corresponding permuted-graph baseline (see “Baseline models” section in Methods) in which protein names were permuted to remove biological meaning from the graph. Bars represent mean test R^2^ across 10 independent training runs (± S.E.M.). The phenotypes evaluated were **(A)** glycated hemoglobin (HbA1c; UKBB Field 30750), **(B)** blood C-reactive protein (CRP) measured from serum biochemistry (UKBB Field 30710), **(C)** pack years smoked (UKBB Field 20161), and **(D)** standing height (UKBB Field 50). When training with the full training set, test R^2^ values obtained from ridge regression (not shown) for HbA1c, blood CRP, pack years smoked, and height were 0.6231, 0.6687, 0.2741, and 0.6721, respectively.

**Supplementary Figure 2.**
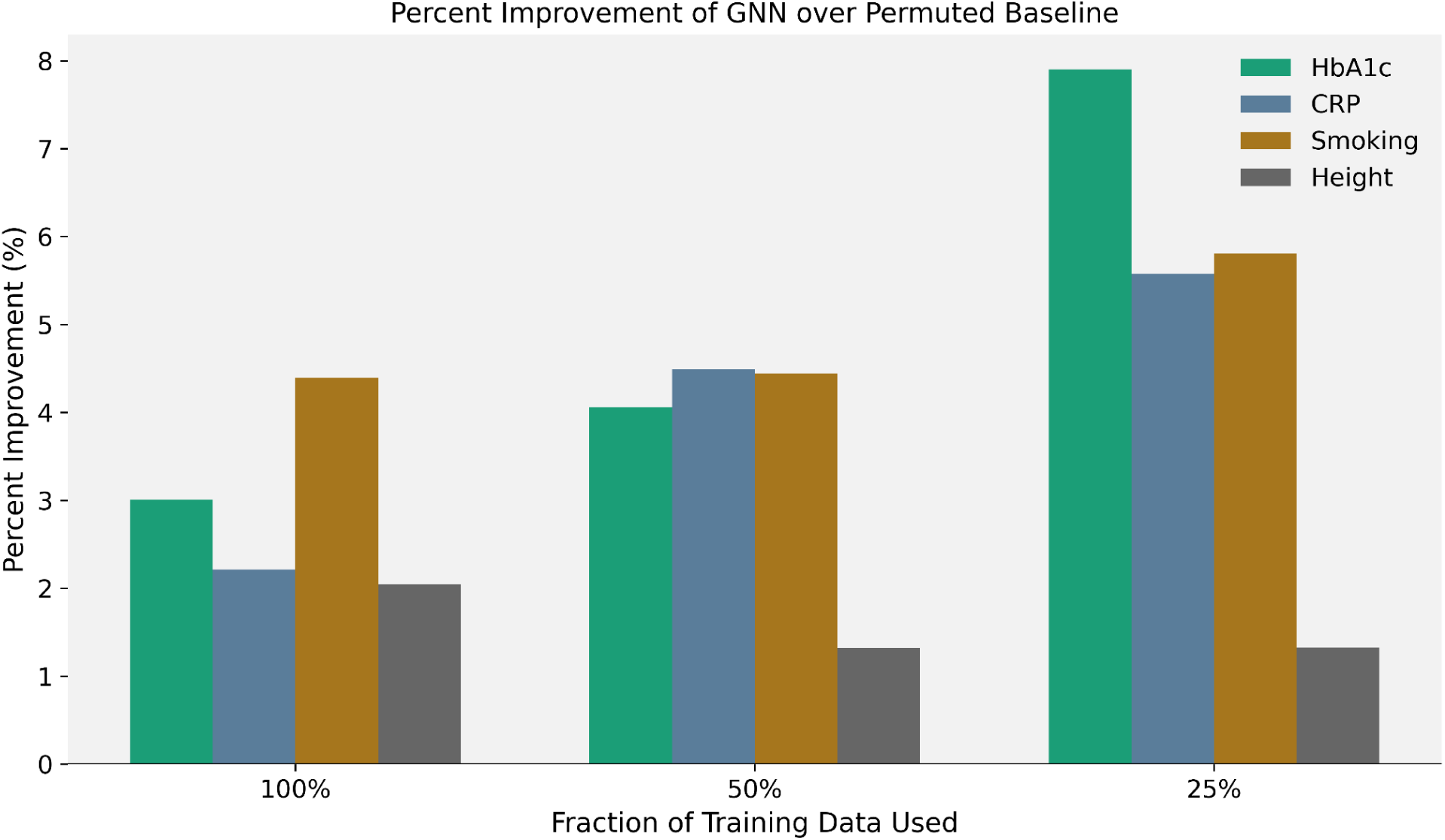
Performance of single-head GNNs compared to permuted GNNs across multiple phenotypes and training data fractions. The single-head GNNs, which operated on the protein graph, were trained on the full training data (100%) and smaller subsets of it (50%, 25%). Each model was compared to a corresponding permuted-graph baseline (see “Baseline models” section in Methods) in which protein names were permuted to remove biological meaning from the graph. Mean percent improvement in test R^2^ (represented by bars) was computed for each model relative to its permuted-graph baseline. The phenotypes evaluated were **(A)** glycated hemoglobin (HbA1c; UKBB Field 30750), **(B)** blood C-reactive protein (CRP) measured from serum biochemistry (UKBB Field 30710), **(C)** pack years smoked (UKBB Field 20161), and **(D)** standing height (UKBB Field 50).

**Supplementary Figure 3.**
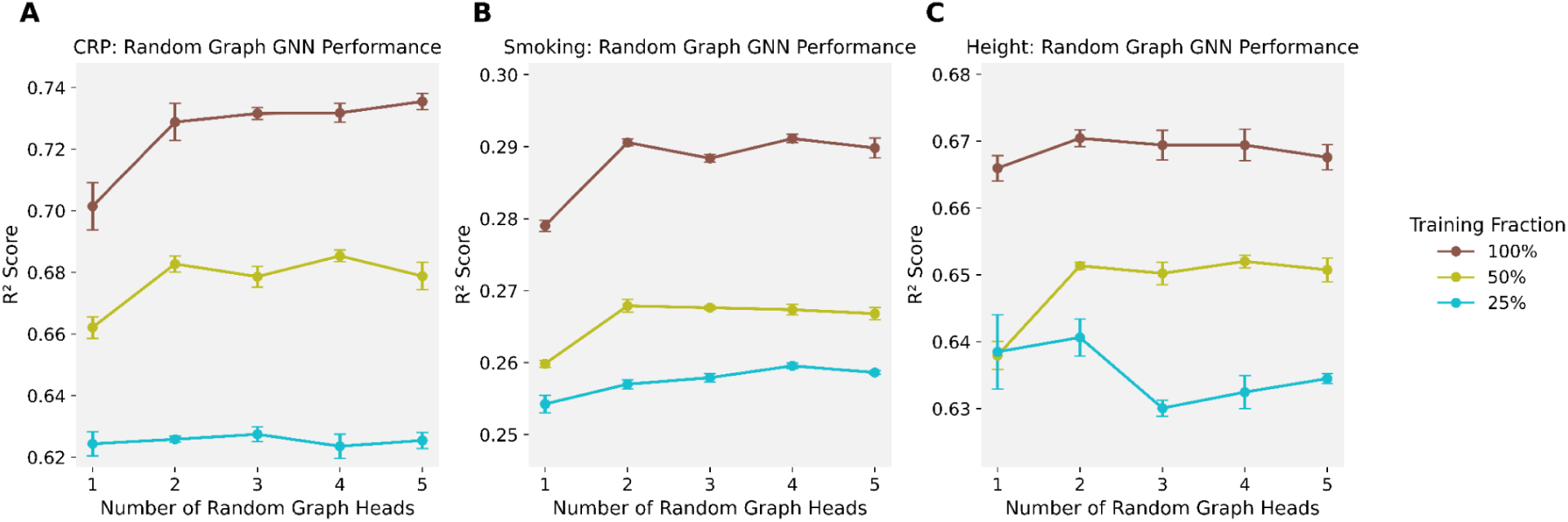
Performance of GNNs trained on random graph structures across additional phenotypes not presented in the main paper. GNNs were trained using randomly generated bipartite graphs containing between one and five random graph heads, with 100%, 50%, and 25% of the training data used for each configuration. Points represent mean test R^2^ across 5 independent training runs (± S.E.M.). The phenotypes evaluated were **(A)** blood C-reactive protein (CRP) measured from serum biochemistry (UKBB Field 30710), **(B)** pack years smoked (UKBB Field 20161), and **(C)** standing height (UKBB Field 50).

## Notes

### Competing Interest Statement

Alexis Battle is a shareholder in Alphabet Inc, and a founder and equity holder of CellCipher, Inc. Joshua M. Popp is an equity holder of CellCipher, Inc.

### Funding Statement

This study was funded by by NIGMS R35GM139580 awarded to Alexis Battle.

### Author Declarations

The ethics committee of the North-West Multi-centre Research Ethics Committee has approved data collection by the UK Biobank. This work was done in concordance with UK Biobank application number 98985, which grants approval for our study.

